# Comparison of Different PCR Methods for the Detection of SARS-CoV-2 RNA in Wastewater Based on the Reported Incidence of COVID-19 in Finland

**DOI:** 10.1101/2023.09.07.23295183

**Authors:** Annika Länsivaara, Kirsi-Maarit Lehto, Rafiqul Hyder, Erja Janhonen, Anssi Lipponen, Annamari Heikinheimo, Tarja Pitkänen, Sami Oikarinen, the WastPan Study Group

## Abstract

The spatial and temporal changes of the COVID-19 pandemic have been monitored with wastewater-based surveillance, which many countries have applied to their national public health monitoring measures. The most commonly used methods for the detection of SARS-CoV-2 in wastewater are RT-qPCR and RT-ddPCR. Previous comparisons of the two methods have produced conflicting results; some found RT-ddPCR to be more sensitive, one found RT-qPCR to be more sensitive, and others found them to be equal in sensitivity. This research was conducted to further study these two methods as well as two different RNA extraction methodologies and gene assays for the detection of SARS-CoV-2 in wastewater. We compared two RT-qPCR kits and RT-ddPCR based on sensitivity, variability, and the correlation of SARS-CoV-2 gene copy numbers in wastewater with the incidence of COVID-19. Our results indicate that the most sensitive and low-variance method to detect SARS-CoV-2 in wastewater was RT-ddPCR. However, we obtained the best correlation between COVID-19 incidence and SARS-CoV-2 gene copy number in wastewater using RT-qPCR (CC = 0.697, p < 0.001). We found a significant difference in sensitivity between the two RT-qPCR kits, one having a significantly lower limit of detection and a higher percentage of positive samples than the other. Furthermore, the CDC N1 primers and probe were the most sensitive for both RT-qPCR kits, while there was no significant difference between the tested gene targets using RT-ddPCR. For the most sensitive RT-qPCR, the use of different RNA extraction kits affected the result. All methods showed a trend between COVID-19 incidence and SARS-CoV-2 gene copy numbers in wastewater. In addition, we tested an isothermal amplification method for the detection of SARS-CoV-2 RNA in wastewater. It proved to be a viable option if results are expected quickly, resources are limited, and presence–absence information is sufficient.

**Highlights:**

- Using different RNA extraction kits, detection kits, and gene assays to detect SARS-CoV-2 in wastewater produces differing results.
- SARS-CoV-2 gene copies in wastewater correlate with the reported incidence of COVID-19 in Finland.
- RT-ddPCR was the most sensitive and repeatable method to detect SARS-CoV-2 in wastewater, whereas RT-qPCR had the best correlation to the incidence of COVID-19.
- RT-SIBA is a viable option for the detection of SARS-CoV-2 in wastewater in low-resource settings.
- All methods have high result variability when the amount of SARS-CoV-2 in wastewater is low.

**Graphical abstract:** Created with BioRender.com

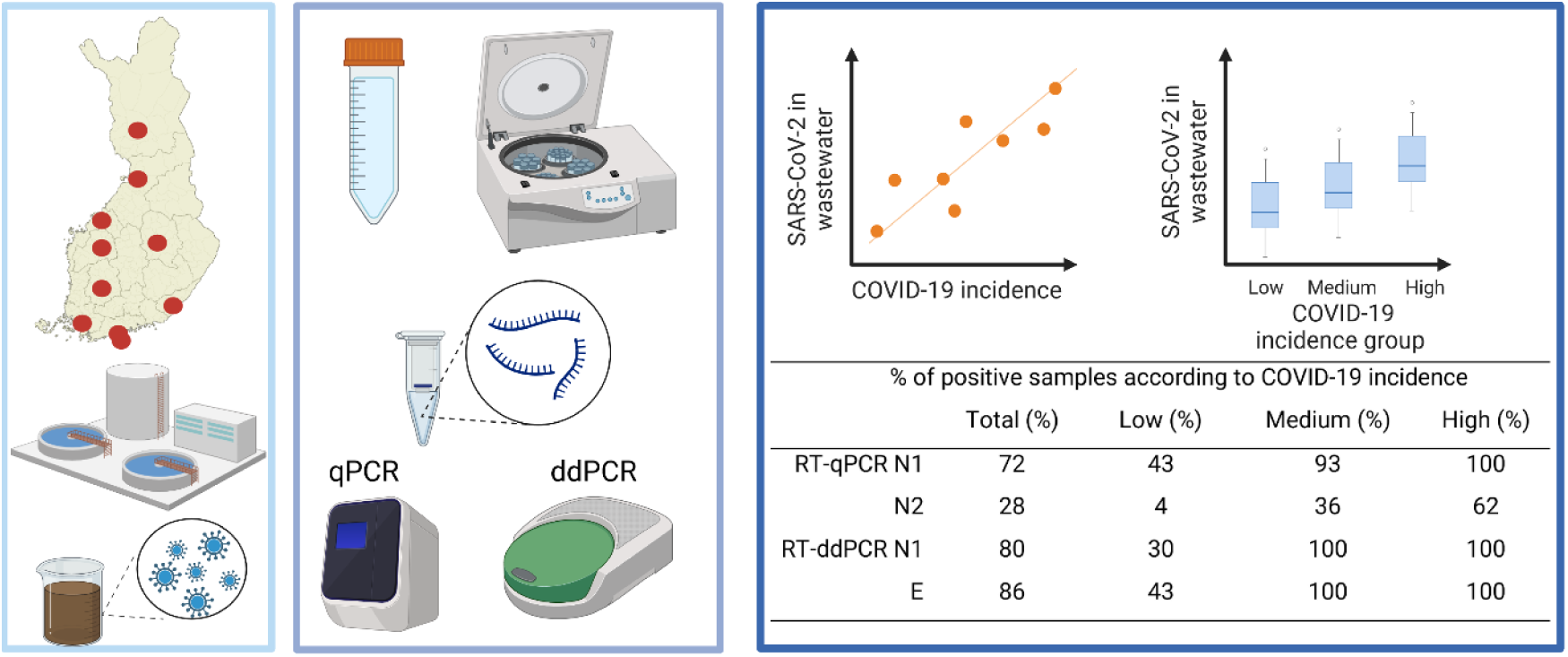

## 1. Introduction

Wastewater reflects the circulation of microbes in the population in certain sewerage network areas and can be used to evaluate the circulation of various pathogens (Mao et al., 2020). SARS-CoV-2 RNA is excreted in the feces of infected individuals (Cerrada-Romero et al., 2022), and the spatial and temporal changes of the COVID-19 pandemic can therefore be studied from wastewater (Ahmed et al., 2020; De Freitas Bueno et al., 2022; Gonzalez et al., 2020; Randazzo et al., 2020). The need for surveillance of SARS-CoV-2 in wastewater has been demonstrated by the fact that the estimated spread of COVID-19 based on wastewater surveillance has been much higher than would be expected based on clinical cases showing the actual spread of the virus (Wu et al., 2020). In addition, it has been shown that wastewater monitoring of SARS-CoV-2 can be used to estimate new hospital and intensive care unit admissions 2–8 days ahead of time (Galani et al., 2022). Furthermore, wastewater provides an easy and cost-efficient pooled sample matrix. A national wastewater surveillance system for SARS-CoV-2 has been implemented in several countries, the information from which is utilized in public health decision making. The Centers for Disease Control and Prevention (CDC) has started national wastewater surveillance in the US (Keshaviah et al., 2021), and the European Commission has called for the systematic surveillance of SARS-CoV-2 in wastewater in the EU (Agrawal et al., 2021). Furthermore, the European Commission has suggested that member states start surveilling influenza A and poliovirus from wastewater (EU, 2022). For the surveillance to be reliable, the use of a suitable method and careful optimization and validation of the method is necessary.

For the wastewater surveillance of SARS-CoV-2, both RT-qPCR and RT-ddPCR are used. To date, comparisons between the two methods have been made regarding SARS-CoV-2 wastewater surveillance, although they have produced conflicting results. One study conducted during low COVID-19 incidence showed ddPCR to be more prone to inhibitors than RT-qPCR, to have a higher limit of detection (LOD), and to estimate lower gene copy numbers (GCs) of the virus (D’Aoust et al., 2021). In contrast, Ahmed et al. (2022b) discovered ddPCR to have a higher positivity rate and LOD than qPCR. Similarly, Ciesielski et al. (2021) noted RT-ddPCR to be more sensitive and to have a lower LOD than qPCR. The higher detection rate, sensitivity, and precision of ddPCR were also noted by Flood et al. (2021). Lucansky et al. (2023) discovered ddPCR to be more specific and sensitive than qPCR. On the other hand, Boogaerts et al. (2021) found ddPCR and qPCR to be comparable in sensitivity. Previous studies have also noted differences in results when using different RNA extraction methods (O’Brien et al., 2021; Zheng et al., 2022) and target gene assays (Ahmed et al., 2022b; Barua et al., 2022; Flood et al., 2021). Currently, in the spring of 2023, the incidence of COVID-19 is on a downward trend (WHO, 2023). As the amount of SARS-CoV-2 in wastewater declines, the need for a sensitive detection method increases. As previous results on the performance of PCR-based detection methods have been conflicting, further research is needed.

RT-qPCR and RT-ddPCR each require extensive resources and expertise. In addition to identifying the most sensitive method to detect SARS-CoV-2 in wastewater, there is also a need for methods that require fewer resources and are easier to use. One option for fast and resource-saving analysis of SARS-CoV-2 in wastewater is reverse transcription strand invasion based amplification (RT-SIBA), a qualitative method based on the isothermal amplification of nucleic acids. Compared to PCR, RT-SIBA is faster, and the instruments required are less complex than they are for methods that require thermal cycling. It has previously been used to detect viruses, such as respiratory syncytial virus (RSV), influenza type A and B viruses, and rhinovirus, in clinical specimens (Eboigbodin et al., 2016, 2017; Kainulainen et al., 2019). According to our knowledge, there have been no studies of RT-SIBA to detect SARS-CoV-2 RNA in wastewater, and its use in the detection of SARS-CoV-2 for diagnostic use has been reported once (Rosenstierne et al., 2021). Previously, one isothermal amplification method, loop-mediated isothermal amplification (RT-LAMP), was used to detect SARS-CoV-2 in wastewater, but the method was shown to be 20 times less sensitive than RT-ddPCR (Bivins et al., 2022).

In this study, we compare the sensitivity and repeatability of various SARS-CoV-2 detection methods, including two RNA extraction methods, two RT-qPCRs kits, and the RT-ddPCR method, using different target genes. Furthermore, the SARS-CoV-2 GCs in wastewater detected by each method are compared to COVID-19 incidence. Samples collected during low incidence of COVID-19 are used in the study to test the methods’ performance using samples with low GC numbers of the target. The study also tests RT-SIBA for the qualitative detection, presence–absence information, of SARS-CoV-2 in wastewater. The sample processing is depicted in Figure 1.

**Figure 1.**
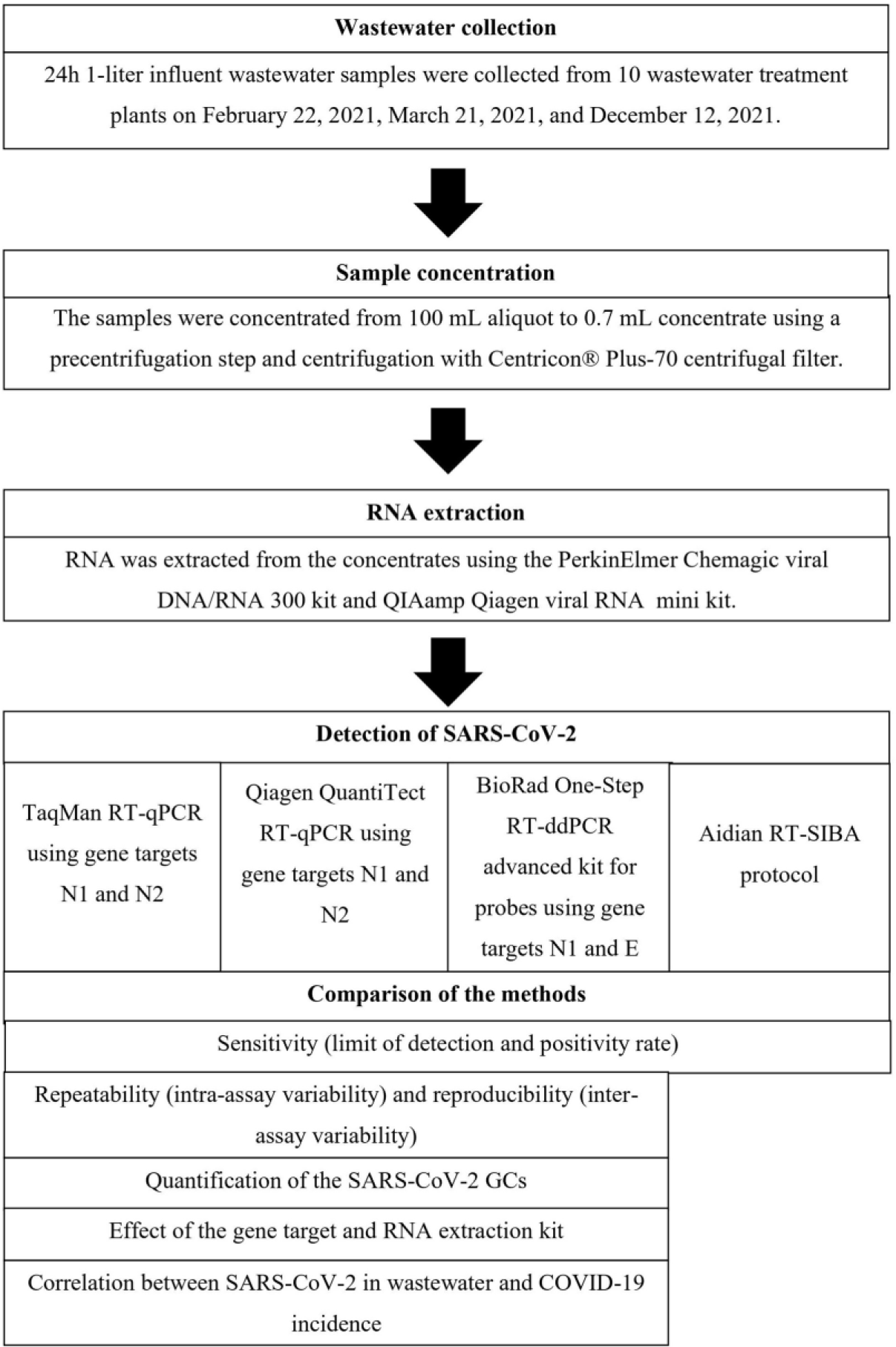
Sample processing flowchart. First, wastewater samples were collected and concentrated and RNA was extracted. Next, SARS-CoV-2 was detected using TaqMan Fast Virus 1-Step RT-qPCR, Qiagen QuantiTect Probe RT-qPCR, Bio Rad One-Step RT-ddPCR, and Aidian SIBA protocol. All the methods were then compared based on their sensitivity. In addition, the quantitative methods (two RT-QPCR methods and the RT-ddPCR method) were compared based on the quantification of SARS-CoV-2 GCs, intra- and inter-assay variability, effect of the gene target and RNA extraction method, and the correlation between SARS-CoV-2 GC in wastewater and COVID-19 incidence.

## 2. Material and Methods

### 2.1 Wastewater Sampling

The samples used in this study are a part of the WastPan project conducted in collaboration with Tampere University, the Finnish Institute for Health and Welfare (THL), and the University of Helsinki between 2020 and 2023. The project aims to develop tools for the wastewater-based surveillance of pathogens and antimicrobial resistance genes. This study includes ten wastewater treatment plants (WWTPs) located in Espoo (Suomenoja WWTP, 390,000 inhabitants), Helsinki (Viikinmäki WWTP, 860,000 inhabitants), Kuopio (Lehtoniemi WWTP, 91,000 inhabitants), Lappeenranta (Toikansuo WWTP, 63,000 inhabitants), Oulu (Taskila WWTP, 200,000 inhabitants), Pietarsaari (Alheda WWTP, 31,000 inhabitants), Rovaniemi (Alakorkalo WWTP, 55,000 inhabitants), Seinäjoki (Seinäjoen keskuspuhdistamo WWTP, 55,000 inhabitants), Tampere (Viinikanlahti WWTP, 200,000 inhabitants), and Turku (Kakolanmäki WWTP, 300,000 inhabitants). The samples used in this article were collected on February 22, 2021; March 21, 2021; and December 12, 2021. In addition, samples were collected from Espoo and Helsinki on May 17, 2021. From 24-h composite samples of influent untreated wastewater, a fraction of 1 L was shipped in cool boxes within 24–48 hours of sampling. After arrival, the samples were frozen and kept in a −80 ⁰C freezer before analysis.

The performance of the methods was tested using samples collected at various COVID-19 incidences in the WWTP areas. The wastewater samples were classified into three groups according to the incidence of COVID-19, based on the reported cases in the Finnish National Infectious Disease Register by the Finnish National Institute for Health and Welfare. Incidence was measured in terms of COVID-19 cases per 100,000 inhabitants over a 7-day period of the sampling week in the city in which the WWTP was located (incidence = cases / population of the WWTP city * 100,000). The low-incidence group (N = 12) included samples collected at a time with less than 50 cases, the medium group 50–200 cases (N = 9), and the high group over 200 cases (N = 9) of COVID-19 per 100,000 inhabitants per week. The low-incidence group included three samples for which the incidence of COVID-19 was zero.

### 2.2 Concentration of Wastewater Samples

Frozen samples were thawed in a refrigerator and then concentrated immediately, according to the method described by Hokajärvi et al. (2021). First, a 100 mL aliquot of the 1 L original sample was melted in the refrigerator for the analysis. Immediately after the sample had melted, interfering particles, such as debris, were removed with centrifugation at 4654 g for 30 min without brake. The supernatant was then concentrated using a Centricon^®^ Plus-70 centrifugal ultrafilter with a cut-off of 100 kDa (Millipore, Cork, Ireland) and centrifuged at 3500 g for 15 min. The concentrate was collected by centrifugation at 1000 g for 2 min.

### 2.3 RNA Extraction

RNA extraction was performed immediately after concentration. All samples were extracted with a PerkinElmer Chemagic Viral DNA/RNA 300 (Wallac Oy, Turku, Finland), and the samples collected in February, March, and May 2021 were also extracted using a Qiagen QIAamp Viral RNA mini kit (Qiagen, Hilden, Germany). Both extractions were done according to manufacturers’ protocols. The sample volume was 300 µL for the Chemagic RNA extraction kit and 140 µL for the QIAamp RNA extraction kit. The elution volume was 100 µL for the Chemagic extraction and 60 µL for the QIAamp extraction. The samples were each extracted twice in the same extraction run to produce enough eluate for the method comparisons. The two eluates of the same sample were then mixed into one sample, which was used for all the detection methods. All the detection methods were performed from the same sample at the same time to enable equal comparison.

### 2.4 SARS-CoV-2 Synthetic RNA Control

Ten-fold standard dilution series (1–10,000 copies/µL) of SARS-CoV-2 synthetic RNA control (Codex DNA, CA, USA) was run on each qPCR plate as a positive control and to quantify the SARS-CoV-2 GCs. An RNA control was also used to calculate the LOD for each method.

### 2.5 RT-qPCR

RT-qPCR was performed using two different RT-qPCR kits, TaqMan Fast Virus 1-Step Mastermix (TaqMan RT-qPCR; Applied Biosystems by Thermo Fisher Scientific, Vilnius, Lithuania) and Qiagen QuantiTect Probe RT-PCR (QuantiTect RT-qPCR; Qiagen, Hilden, Germany), and the CDC N1 and N2 primer-probe sets. The Sarbeco E-gene primer-probe set (Corman et al., 2020) was also tested during the optimization of the RT-qPCR methods; we noted that most samples were non-detects, and thus, they were rejected from the study. The sequences of the primers and probes as well as the cycling conditions for all the RT-qPCR reactions are shown in the supplementary material (Tables S.1, S.2 and S.3). The specificity and sensitivity of the primers and probes were validated with the Quality Control for Molecular Diagnostics (QCMD) panel. Cross-reactivity to endemic coronaviruses 229E and OC43 was also tested using the panel, and no cross-reactivity was found. The TaqMan RT-qPCR was performed according to the manufacturer’s protocol using a total volume of 25 µL. The reaction mixture for the TaqMan N1 and N2 assays included 6.25 µL of the TaqMan Fast Virus 1-step mastermix, 200 nM forward primer, 200 nM reverse primer, 200 nM probe, and 5 µL template. The QuantiTect RT-qPCR had been previously optimized for clinical samples using a total volume of 10 µL and a template volume of 2 µL; initially, these volumes were used. The reaction mixture for the QuantiTect N1 gene assay included 5 µL of the QuantiTect RT mastermix, 900 nM forward primer, 900 nM reverse primer, 200 nM probe, 1 µL QuantiTect RT mix, and 2 µL template. The reaction mixture for the QuantiTect N2 gene assay included 5 µL of the QuantiTect RT mastermix, 300 nM forward primer, 900 nM reverse primer, 200 nM probe, 1 µL QuantiTect RT mix, and 2 µL template. In addition, the December 2021 samples were analyzed with the QuantiTect RT-qPCR assay using the manufacturer-recommended total volume of 25 µL and template volume of 5 µL while using the same primer and probe concentrations to evaluate the effect of the template volume. Negative control for RNA extraction and qPCR negative and positive controls were run on each plate. The samples were run in triplicate. Repeatability (intra-assay variation) was analyzed among the triplicates, and reproducibility (inter-assay variation) was determined using three different mastermix reactions, for a total of nine repetitions. The runs were performed on the Applied Biosystems QuantStudio 5 Real-Time PCR System.

The 95% LOD was calculated using regression probit analysis (Stokdyk et al., 2016). The first four different concentrations of the SARS-CoV-2 synthetic RNA control (Codex DNA, CA, USA) were replicated 12 times, after which the proportion of positive reactions (probability of detection) was calculated. The probability of detection was transformed into probability units by the inverse of the normal cumulative distribution, and the concentrations were then transformed into base 10 logarithm. The probability units were plotted against the base 10 logarithms of the concentrations. Finally, the 95% limit of detection was calculated by solving the regression equation, where y = probability unit = 1.64 (probability unit 1.64 equites to 95% probability). The efficiency, standard curve slope, standard curve intercept, and R² of each RT-qPCR assay are reported in the supplementary material (Table S.4).

### 2.6 RT-ddPCR

RT-ddPCR was performed using the Bio Rad One-Step RT-ddPCR Advanced Kit for Probes (RT-ddPCR; Bio Rad, Hercules, CA, USA). Preliminary testing of the CDC N1, N2, and N3 primer-probe sets (Lu et al., 2020) and the Sarbeco E-gene primer-probe set (Corman et al., 2020) showed that the CDC N1 and the Sarbeco E-gene primer-probe sets were the most sensitive (data not shown). The sequences of the primers and probes as well as the cycling conditions for all the RT-ddPCR reactions are shown in the supplementary material (Tables S.1 and S.5). RT-ddPCR was performed according to manufacturer’s protocol using a template volume of 4 µL and a total volume of 20 µL. The reaction mixture for the N1 and E assays included 5.5 µL of supermix, 100 nM forward primer, 100 nM reverse primer, 25 nM probe, 1.1 µL diothiothreitol, 2.2 µL RT enzyme, and 4 µL template. A negative and a positive control were used in each run. All samples were run in duplicate, apart from the reproducibility analyses, which were run in triplicate with three different master mixes, for a total of nine runs. Droplets were generated using a Bio Rad QX200 droplet generator and read after the PCR run using a Bio Rad QX200 droplet reader.

### 2.7 RT-SIBA

RT-SIBA was performed for the wastewater samples collected in December 2021 using the Aidian SIBA protocol for SARS-CoV-2. In addition, the sensitivity of the method was determined using the SARS-CoV-2 synthetic RNA control. First, a master mix was prepared by combining 15.2 µL of the RT-SIBA A-MIX, 15.2 µL of the RT-SIBA B-MIX, and 7.6 µL of the Oligomix 1 High SYBR 1.0. Then, 0.43 µL of magnesium acetate (Sigma-Aldrich) was added to the RNA template, after which 5 µL of the RNA template was added to the master mix. A positive RNA control, a negative control for RNA extraction, and a RT-SIBA negative control were run on each plate. The SIBA reaction was run on the BioRad CFX96 Real-Time PCR detection system. The cycling conditions are presented in the supplementary material (Table S.6). All the samples were run in triplicate, apart from the repeatability analyses, which were run in triplicate at three different times, for a total of nine runs.

### 2.8 Statistical Analysis

All statistical analyses were performed on IBM^®^ SPSS^®^ software version 28.0.1.0 (142). Statistical significance was tested using p-values calculated with the Kruskal–Wallis H test. Correlations between SARS-CoV-2 GC numbers in wastewater and the incidence of COVID-19 cases were evaluated using the Kendall rank correlation coefficient.

## 3. Results

### 3.1 RT-qPCR

The 95% LOD for the RT-qPCR was determined for each of the assays using the standard series of SARS-CoV-2 synthetic RNA control and the method described by Stokdyk et al. (2016). The LOD was 18.4 GC/µL for the TaqMan N1 assay, 19.9 GC/µL for the TaqMan N2 assay, 77.6 GC/µL for the QuantiTect N1 assay, and 80.7 GC/µL for the QuantiTect N2 assay. TaqMan RT-qPCR detected SARS-CoV-2 in 72% of the wastewater samples, and QuantiTect RT-qPCR did so in only 20% of the samples (Table 1).

**Table 1.** The proportion of positive samples (%) indicated by the test methods and gene targets in wastewater samples in different incidence sample groups. The low category includes three samples that were collected when the incidence of COVID-19 was zero. Low incidence N = 12, medium incidence N = 9, high incidence N = 9.

|  | Incidence of COVID-19 |  |  |  |
| --- | --- | --- | --- | --- |
|  | Total (%) | Low (%) | Medium (%) | High (%) |
| TaqMan RT-qPCR N1 | 72 | 43 | 93 | 100 |
| N2 | 28 | 4 | 36 | 62 |
| QuantiTect RT-qPCR N1 | 20 | 9 | 21 | 38 |
| N2 | 6 | 0 | 0 | 23 |
| RT-ddPCR N1 | 80 | 30 | 100 | 100 |
| E | 86 | 43 | 100 | 100 |

The sensitivity of the N1 and N2 gene regions were compared. For both RT-qPCR methods, the N1 gene region was more sensitive. The LOD was lower for the N1 assay than for the N2 assay for both RT-qPCR kits. For TaqMan, 72% of the samples were positive with the N1 region, whereas only 28% were positive with the N2 region. For QuantiTect, using the N1 gene region resulted in 20% of the samples being positive, and 6% with the N2 region. The N1 gene region was more sensitive in all incidence sample groups (Table 1). There was no statistically significant difference in the GC numbers detected by the two gene targets within the RT-qPCR assays.

The effect of the RNA extraction method was also studied by analyzing the February and March samples with the RNA extraction kits manufactured by Chemagic and Qiagen. With the TaqMan RT-qPCR, 40% of the samples extracted with Chemagic were positive and 35% of those extracted with Qiagen were positive. The GC numbers detected from the samples extracted with Chemagic were slightly higher than for those extracted with Qiagen (Fig. 2). The result was statistically significant (p < 0.001). With the QuantiTect RT-qPCR, 7.5% of the samples were positive using both extraction kits. There was no statistically significant difference in the GC numbers detected by the QuantiTect RT-qPCR using the two extraction kits.

**Figure 2.**
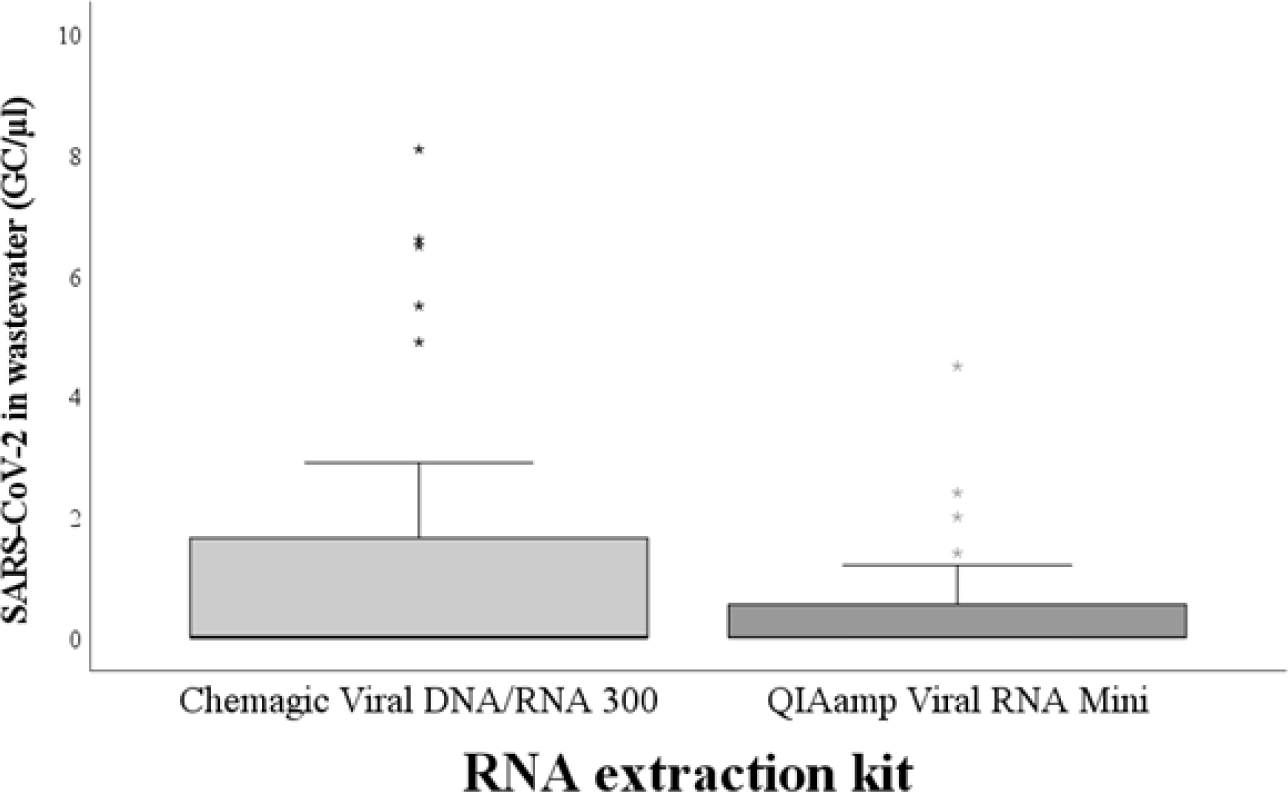
The difference in SARS-CoV-2 GC numbers per µL of sample between the PerkinElmer Chemagic Viral DNA/RNA 300 and Qiagen QIAamp Viral RNA mini kit. The difference was observed using the TaqMan Fast Virus 1-step RT-qPCR (p = 0.035).

To assess the repeatability (intra-assay variability) and reproducibility (inter-assay variability) of the most sensitive RT-qPCR method, two samples were analyzed using nine replicates. The samples were collected during low incidence of COVID-19 (20 cases per 100,000 persons and 25 cases per 100,000 persons) so that variability could be assessed from samples that contained a low number of SARS-CoV-2 GCs. The results of the TaqMan RT-qPCR showed high variability (Table 2).

**Table 2.**
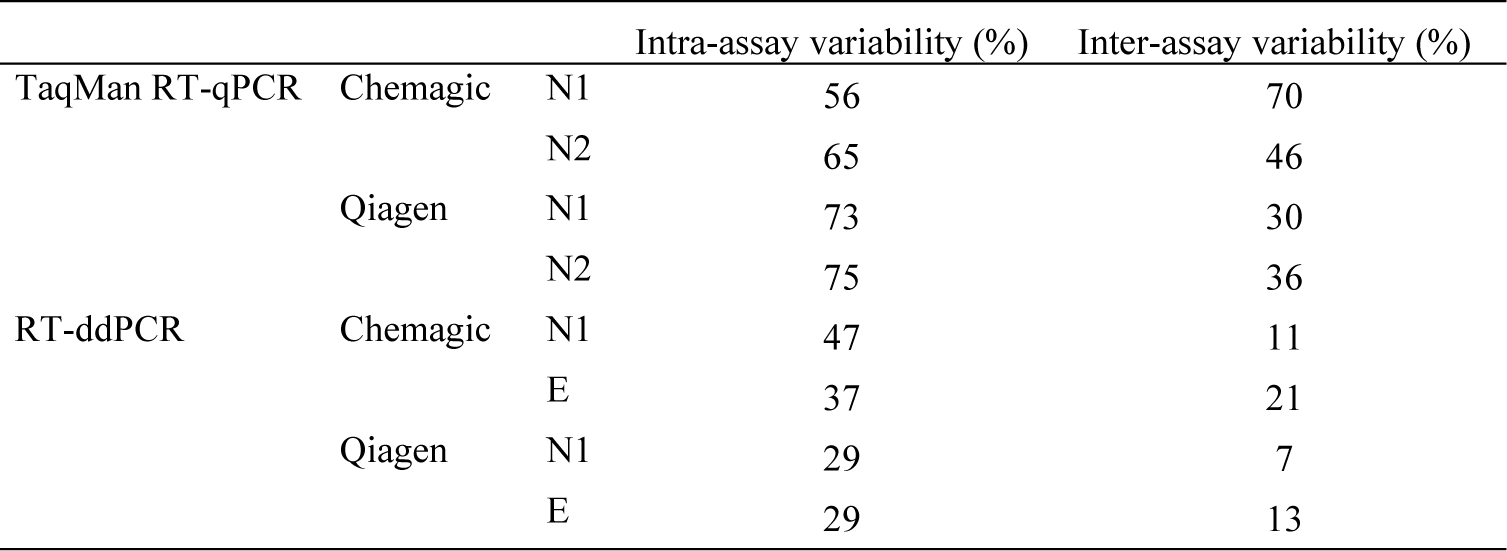
The variability of the TaqMan RT-qPCR and RT-ddPCR presented separately for the Chemagic ja Qiagen RNA extraction kits and N1, N2, and E assays.

For the QuantiTect RT-qPCR, the template volume used was initially 2 µL/reaction. In addition, the manufacturer recommended 5 µL of template per reaction, which was also the template volume for the TaqMan RT-qPCR. The effect of the template volume used was studied using the samples collected in December 2021. The 2 µL/reaction template volume resulted in 40% of the samples being positive and an average of 1.1 SARS-CoV-2 gene copies/µL. The 5 µL/reaction template volume resulted in 10% of the samples being positive and an average of 0.2 SARS-CoV-2 copies/µL, showing that increasing the template volume negatively affected the detection of SARS-CoV-2.

A positive association was seen between the incidence of COVID-19 and the percentage of SARS-CoV-2-positive wastewater samples (Table 1). With the TaqMan RT-qPCR assay, 43% of the low-, 93% of the medium-, and 100% of the high-incidence samples were positive. The same trend was seen in the QuantiTect RT-qPCR assay, with 9% of the low-, 21% of the medium-, and 38% of the high-incidence samples being positive. In addition, a positive association was found between the incidence of COVID-19 and the GC numbers of SARS-CoV-2 in wastewater (Fig. 3). For the TaqMan RT-qPCR assay, the mean GC/µL was 0.6 (0– 14.9) for the low-, 3.0 (0–24.9) for the medium-, and 7.0 (0–28.7) for the high-incidence sample groups. For the QuantiTect RT-qPCR assay, the mean GC/µL was 0.0 (0–0.1) for the low-, 0.1 (0–2.7) for the medium-, and 0.4 (0–8.9) for the high-incidence sample groups. The differences between the GCs of different incidence groups were statistically significant for the TaqMan RT-qPCR (p < 0.001) but not for the QuantiTect RT-qPCR. Notably, in almost all the samples of our study, the calculated GCs were under the LOD. Furthermore, the Kendall rank correlation coefficient (CC) was calculated for the SARS-CoV-2 GCs in wastewater detected by each method and the incidence of COVID-19 (Fig. 4). For the TaqMan RT-qPCR, there was a positive correlation, at CC = 0.697 (p < 0.001). As most of the samples were identified as negative by the QuantiTect RT-qPCR, the correlation was not as clear (CC = 0.351, p = 0.018).

**Figure 3.**
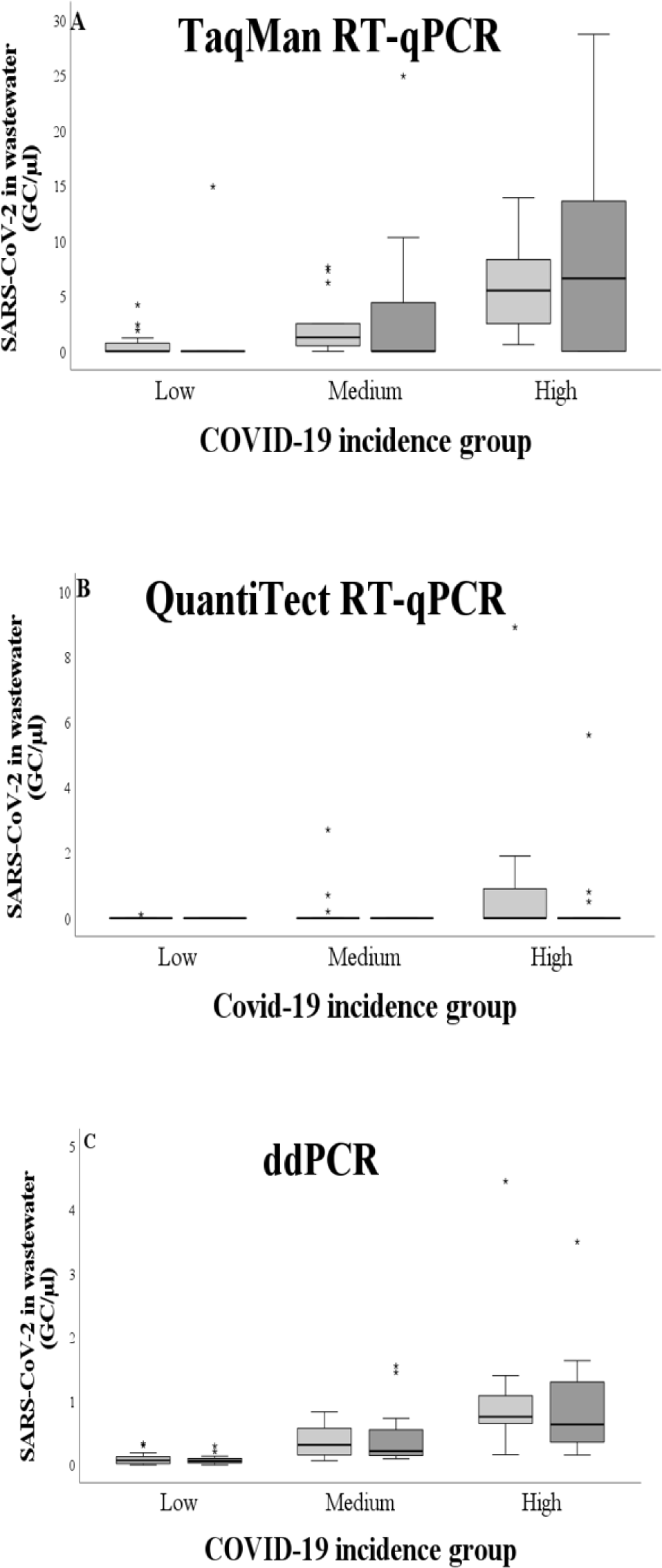
The gene copy numbers of SARS-CoV-2 in different COVID-19 incidence groups with both RT-qPCR kits, RT-ddPCR, and gene targets. a) shows the dataset of TaqMan RT-qPCR N1 (light gray) and N2 assay (dark gray), b) the dataset of QuantiTect RT-qPCR N1 (light gray) and N2 assay (dark gray), and c) the dataset of RT-ddPCR N1 (light gray) and E assay (dark gray).

**Figure 4.**
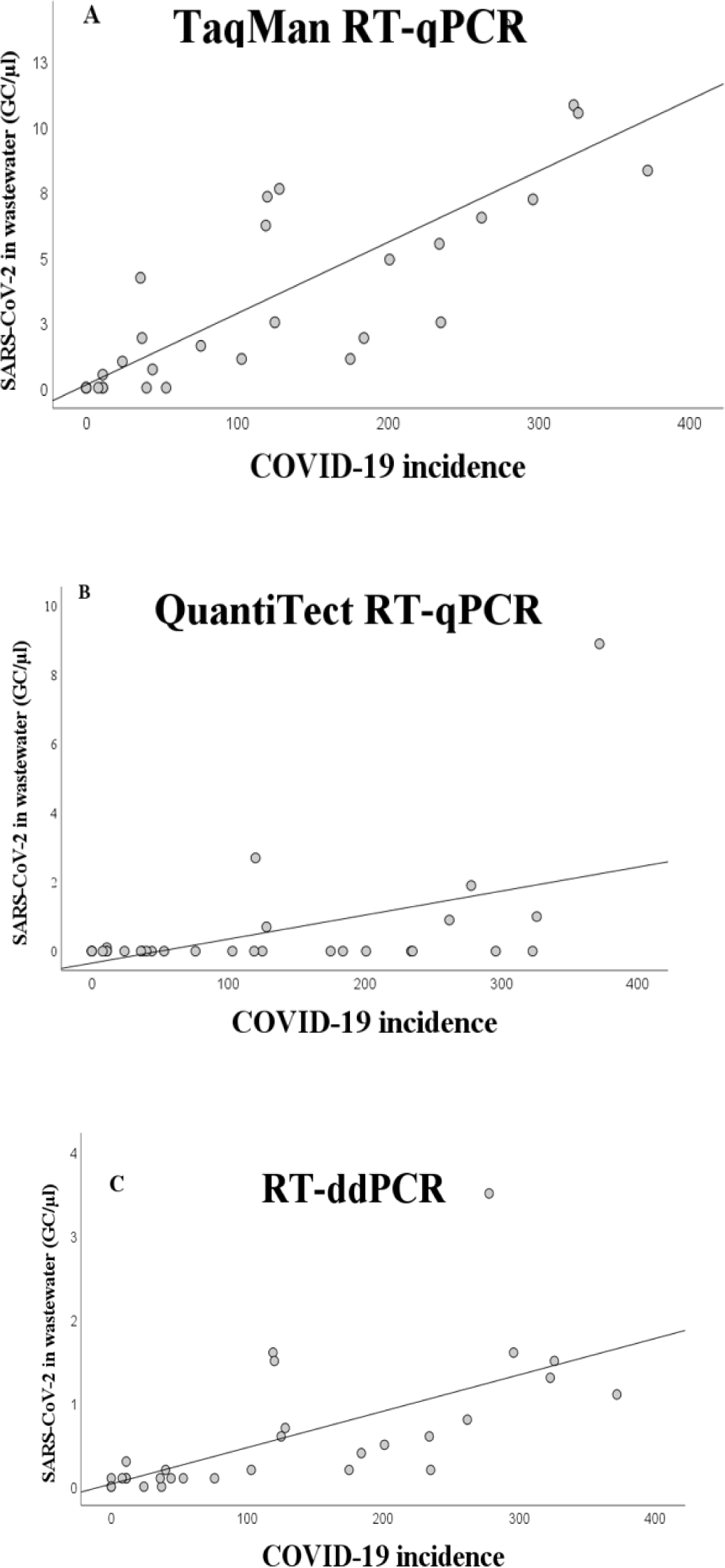
Correlation between the SARS-CoV-2 GC numbers in wastewater and COVID-19 incidence. a) For the TaqMan RT-qPCR, the Kendall rank correlation coefficient was 0.697 (p < 0.001); b) for the QuantiTect RT-qPCR, it was 0.351 (p = 0.018); and c) for the RT-ddPCR, it was 0.629 (p < 0.001). The correlation was calculated using the most sensitive gene target of each method, meaning that the TaqMan and QuantiTect RT-qPCR gene copies included the test results detected with the N1 assay and the RT-ddPCR gene copies the E assay.

### 3.4 RT-ddPCR

The detection limit for RT-ddPCR was determined to be 0.06 GC/µL using the standard series of SARS-CoV-2 synthetic RNA control. Out of all the wastewater samples, 86% were positive when analyzed with RT-ddPCR.

The effects of the gene target and the RNA extraction method used were studied. The E gene assay detected a slightly higher number of positive samples (86%) than the N1 gene assay (80%) (Table 1). There was no statistically significant difference between the SARS-CoV-2 GCs of the two tested gene targets. Furthermore, there was no statistically significant difference in GCs detected between the Chemagic and Qiagen extraction kits.

To assess the repeatability (intra-assay variability) and reproducibility (inter-assay variability) of the detection, two wastewater samples were analyzed using nine replicates. The samples were collected during low incidence of COVID-19 (20 cases per 100,000 persons and 25 cases per 100,000 persons) so that variability could be assessed from samples that contained a low amount of SARS-CoV-2. The results of RT-ddPCR showed high variability (Table 2).

A positive association was found between the incidence of COVID-19 and the percentage of SARS-CoV-2-positive wastewater samples (Table 1). From the low-incidence group of samples, 43% were positive, whereas all samples from the medium- and high-incidence groups were positive (Table 1). Furthermore, a positive association was observed between the incidence of COVID-19 and the SARS-CoV-2 GCs in wastewater (Fig. 3). The mean GC number was 0.08 (0–0.3) for the low-, 0.4 (0.1–1.6) for the medium-, and 1.0 (0.2–4.5) for the high-incidence groups. The results were statistically significant (p < 0.001). Furthermore, the SARS-CoV-2 GC numbers in wastewater correlated with the incidence of COVID-19 (CC = 0.629, p < 0.001) (Fig. 4).

### 3.5 Correlation Between Methods

The correlation between the methods was investigated using wastewater results. A positive correlation was observed between the TaqMan RT-qPCR and RT-ddPCR assays (CC = 0.783, p < 0.001) (Fig. 5). However, the quantification of SARS-CoV-2 GC numbers in the wastewater samples was much higher with the TaqMan RT-qPCR than with the RT-ddPCR method. No correlation was found between the QuantiTect RT-qPCR assay and other methods due to the low positivity rate of the QuantiTect RT-qPCR assay.

**Figure 5.**
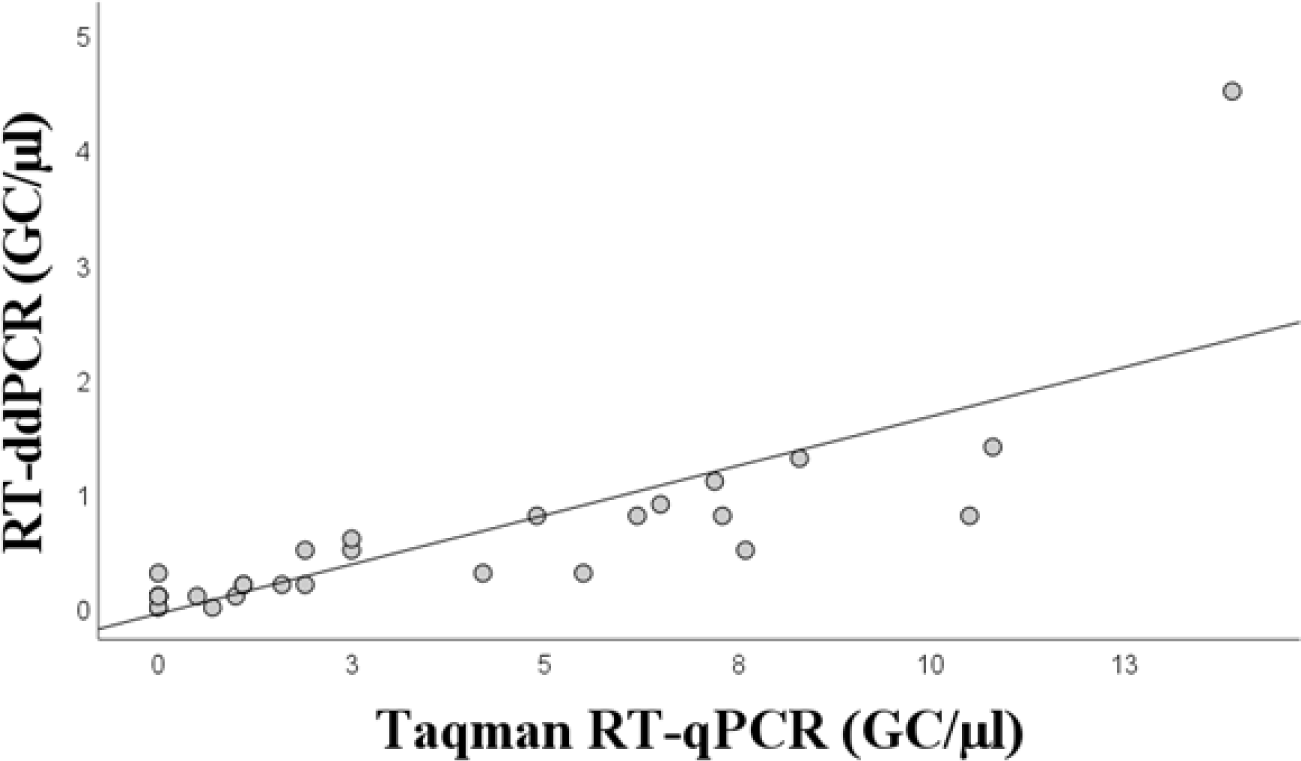
Correlation between the SARS-CoV-2 gene copy numbers in wastewater detected by TaqMan RT-qPCR and RT-ddPCR. The Kendall rank correlation coefficient was 0.783 (p < 0.001). The test results analyzed using the N1 assay were included.

### 3.6 RT-SIBA

The detection limit for RT-SIBA was determined to be 23 GC/reaction using the standard series of SARS-CoV-2 synthetic RNA control. On average, 10 GC/µL of the template was detected in 22 minutes, 100 GC/µL in 19 minutes, 1000 GC/µL in 15 minutes, and 10,000 GC/µL in 12 minutes. The wastewater samples collected in December 2021 were analyzed by RT-SIBA assay. All these samples were positive for SARS-CoV-2.

## 4. Discussion

This study was conducted to compare different methodologies in the analysis of SARS-CoV-2 RNA in wastewater. Since many countries have already implemented such analysis in their public health programs, there is a great need for a sensitive and reliable method for the analysis of SARS-CoV-2 in wastewater. Furthermore, the European Commission has recommended that its member states start surveilling other viruses, such as influenza A, from wastewater. SARS-CoV-2 studies on methodology could be used to aid the selection of the method for the surveillance of other viruses. It is critical for the detection method to have high sensitivity for the successful usage of WBS as an early warning tool for pandemics and for the detection of disease outbreaks. Previous studies have provided conflicting information on the sensitivities of RT-qPCR and RT-ddPCR, with one finding RT-qPCR to be more sensitive (D’Aoust et al., 2021), some finding ddPCR to be more sensitive (Ahmed et al., 2022b; Ciesielski et al., 2021; Flood et al., 2021; Lucansky et al., 2023), and similar sensitivities observed between the methods (Barua et al., 2022; Boogaerts et al., 2021), indicating the need for further research on the topic. In this study, we tested two different RT-qPCR kits using the same method to show that significant differences can be found among different kits. Most previous studies have only tested one RT-qPCR kit. Furthermore, RT-qPCR and RT-ddPCR both require a high level of expertise and resources. Since it would also be important to set up possibilities for SARS-CoV-2 surveillance with limited resources, we also tested isothermal RT-SIBA to provide an option for SARS-CoV-2 wastewater analysis in low-resource settings and when presence–absence information on the virus is sufficient.

Based on our study, the most sensitive method for detecting SARS-CoV-2 in wastewater was RT-ddPCR, since its limit of detection was lower than that of RT-qPCR and it had a higher positivity rate. Furthermore, it had a lower variance of results. This is in line with most of the previous studies (Ahmed et al., 2022a, Ahmed et al., 2022b; Ciesielski et al., 2021; Flood et al., 2021; Lucansky et al., 2023). However, another study reported that RT-qPCR was more sensitive in detecting the virus from post-grit solids and primary clarified sludge because of its lower LOD and detected copy numbers (D’Aoust et al., 2021). Some studies have found the methods to be similar in sensitivity (Barua et al., 2022; Boogaerts et al., 2021).

In our study, we found there to be a difference in virus quantification between RT-ddPCR and RT-qPCR methods. According to the manufacturer, the RNA standard for SARS-CoV-2 contained 10,000 GC/µl, but RT-ddPCR was shown to contain only 253 GC/µl. D’aoust et al. (2021) noted that the GCs of the samples were lower according to ddPCR than to qPCR. Another study found the GCs detected by the two methods to be similar (Barua et al., 2022). The GC numbers of the RNA standard might not be completely correct, as RNA degrades rapidly. The standard used in the quantification of the RT-qPCR may affect its results depending on the accuracy of the standard’s reported GCs. Differences in quantification between different methods should be considered when estimating the amount of SARS-CoV-2 in wastewater. However, a linear positive correlation was observed between the results obtained by the TaqMan RT-qPCR and RT-ddPCR. The problem with differences in quantification can be lessened by following the trends and trendlines of SARS-CoV-2 in wastewater.

All tested methods showed a positive association between the incidence of COVID-19 and the amount of SARS-CoV-2 detected in wastewater. The strongest correlation was found using the TaqMan RT-qPCR with the N1 gene as the detection target. The use of the N2 gene resulted in a weaker correlation, possibly due to its lower sensitivity. Although RT-ddPCR was the most sensitive method, its correlation with the incidence of COVID-19 was weaker than that of the TaqMan RT-qPCR N1 gene assay. Still, it should be considered that almost all the samples were under the LOD of RT-qPCR, and thus, the quantification of the copy numbers by RT-qPCR might not be reliable. There was no major difference in the correlation coefficients of RT-ddPCR between the two gene targets. The correlation between the QuantiTect RT-qPCR assays and the incidence of COVID-19 was weak due to low sensitivity. These results show that wastewater surveillance can be used to estimate the circulation of SARS-CoV-2 in a population with optimized PCR-based assays.

Most of the previous methodology articles comparing RT-ddPCR and RT-qPCR only used one RT-qPCR kit. In this study, we wanted to study the differences between kits using the same method. The TaqMan RT-qPCR had a significantly lower LOD (18.4 GC/µL for the N1 assay and 19.9 GC/µL for the N2 assay) than the QuantiTect RT-qPCR (77.6 GC/µL for the N1 assay and 80.7 GC/µL). This shows that the two RT-qPCR kits have significant differences in their sensitivity. This was reflected in the analysis of the samples because the TaqMan RT-qPCR kit had a higher positivity rate (72% N1 gene, 20% N2 gene) than the QuantiTect RT-qPCR (20% N1 gene, 6% N2 gene). These results show that there were significant differences in the performance of RT-qPCR kits even though the methods use the same oligonucleotides. This could partly explain the differences between previous studies, as the studies used different RT-qPCR kits. The difference between kits and assays within one method should always be considered in addition to the differences between RT-qPCR and RT-ddPCR when the results of different studies are compared. Furthermore, it is vital that the methods are described in detail according to the approved guidelines for a reliable comparison of the studies (Bustin et al., 2009).

For RT-ddPCR, the LOD was the same for both target gene assays, and the positivity rate was very similar (86% for the E assay and 80% for the N1 assay). Based on these results, it cannot be evaluated whether one gene assay is better than the other. Previously, N1 was reported to be the most sensitive gene target for RT-ddPCR (Ahmed et al., 2022a), but it is also prone to outliers and correlates poorly with other gene targets (Ho et al., 2022). For both RT-qPCR kits, a significant difference between the two target gene assays was noted. The LOD was lower for the N1 assay than the N2 assay for both TaqMan RT-qPCR and QuantiTect RT-qPCR. Furthermore, the positivity rate was also higher for the N1 assay using both RT-qPCR kits. For TaqMan, the positivity rate was 72% for the N1 assay and 20% for the N2 assay. For QuantiTect, the positivity rate was 20% for the N1 assay and 6% for the N2 assay. The RT-qPCR results are in agreement with previous studies (Ahmed et al., 2022a; Barua et al., 2022). In contrast, Flood et al. (2021) discovered N2 to perform better than N1, N2, and E target genes. They also noted that the E gene resulted in nearly all samples being non-detects when using RT-qPCR. We obtained similar results at the optimization phase of the RT-qPCR methods, and therefore, the E gene was not used in this study. There was no statistically significant difference between the target gene assays in the detected GCs. Even though it can be assumed that a much larger number of positive samples would make the N1 GC average higher, the N2 GCs of the positive samples were higher than the N1 GCs of those samples, making the difference not statistically significant. In a previous study, it was noted that, using RT-qPCR, N2 was more prone to inhibition than N1, while a difference in inhibition was not noted between the gene targets using RT-ddPCR. RT-ddPCR has been noted to be less sensitive to inhibition than RT-qPCR, most likely due to the nanodroplet quantification (Barua et al., 2022). This could explain why studies have not found significant differences in the target gene assays with ddPCR, whereas significant differences have been noted with qPCR.

Furthermore, a difference in the results of the TaqMan RT-qPCR was observed when two different RNA extraction methods were used. Using the Chemagic RNA extraction kit produced higher GCs and positivity rate compared to the QIAamp RNA extraction kit. The Chemagic RNA extraction concentrated samples 3 times and the QIAamp kit did so 2.3 times. Previous studies have also noted differences in the results when using different RNA extraction kits (O’Brien et al., 2021; Zheng et al., 2022). O’Brien et al. (2021) achieved the best result using an RNA extraction kit that had PCR inhibitor removal. Our RNA extraction methodologies did not have inhibitor removal, which could have resulted in the RT-qPCR being negatively affected by inhibition, as we suspected. Zheng et al. (2022) noted that they had significant differences in the sensitivities of the analysis when using two different RNA extraction kits, as one of them was able to detect 1 GC/1 mL of wastewater and the other 10 GC/1 mL of wastewater. These results show that the whole sample processing protocol should be optimized for the sample matrix. It also emphasizes the difficulty of comparing the results of different studies. None of the previous methodology studies used completely the same analysis pipeline, meaning the same RNA extraction kit, RT-qPCR and RT-ddPCR kits, and gene assay. This should always be taken into consideration when different studies are compared. To ensure the quality and comparability of a study, the methods as well as their quality control should be depicted in detail.

Both RT-ddPCR and RT-qPCR had a high variation in the quantification of SARS-CoV-2 GCs in wastewater. In the repeatability and reproducibility test, the lowest intra-assay variability was 29% for RT-ddPCR and 56% for the TaqMan RT-qPCR, and the lowest inter-assay variability was 7% for RT-ddPCR and 30% for the TaqMan RT-qPCR. The variance differed between gene targets and RNA extraction kits, but no clear pattern was found. It is known that variation increases close to the detection limit. This may partly explain the results, since the variability was tested using samples that had a very low number of SARS-CoV-2 GCs. The high variability is likely partly due to wastewater being the sample matrix and to the low GCs of the samples; when using the TaqMan RT-qPCR and an RNA standard of 10,000 GC/µL, we noted the standard deviation to be approximately 15%. Furthermore, one study on SARS-CoV-2 detection in wastewater suggested variability to be most likely affected by the inhibitors present in wastewater (Scott et al., 2023). The variability and poor reproducibility of SARS-CoV-2 wastewater detection was also noted as a common problem in another recent review (Bivins et al., 2021). When interpreting the number of SARS-CoV-2 GCs in wastewater, variation in detection methods should be considered. Particularly during low incidence of virus in the community, the estimation of virus levels in wastewater may be inaccurate.

An isothermal amplification method, RT-SIBA, was tested for the detection of SARS-CoV-2 in wastewater. The sensitivity of RT-SIBA (23 GC/reaction) was similar to that of an RT-SIBA assay used for clinical diagnostics (25 GC/reaction) (Rosenstierne et al., 2021). The RT-LAMP method was also applied for the wastewater analysis of SARS-CoV-2. The assay was less sensitive than RT-SIBA (LOD of 76 N1 GC/reaction) (Bivins et al., 2022). RT-SIBA was used for testing 10 wastewater samples in this study, and all the tested samples were positive. The method was able to provide results quickly. RT-SIBA detected 10 GC/µL of SARS-CoV-2 in 22 minutes. Previously, RT-SIBA was reported to detect 100 GCs of influenza types A and B virus in 15 minutes (Eboigbodin et al., 2016) and 10 GCs of RSV in 20 minutes (Eboigbodin et al., 2017). These results indicate that RT-SIBA is a potential alternative for the detection of SARS-CoV-2 in wastewater. It is a viable option when results are needed quickly and presence–absence information is sufficient. In addition, the method is also easier and cheaper to use than RT-qPCR or RT-ddPCR. It does not require complicated instruments, as the reaction is isothermal (Eboigbodin et al., 2016).

## 5. Conclusion

In October 2022, the European Commission suggested that wastewater be systematically monitored for SARS-CoV-2 and proposed that influenza A and poliovirus also be surveilled from wastewater (EU, 2022). However, wastewater is a complex sample matrix for monitoring SARS-CoV-2, compared to clinical samples. Our study found that the compatibility of all analysis steps, including the RNA extraction method, RT-PCR kit, and gene assay used, as well as quantitative control, influence the performance of the tests; therefore, all analysis steps must be optimized for wastewater samples. Currently, SARS-CoV-2 is monitored mostly with RT-qPCR assays. Yet, our study, as well as most previous studies, showed RT-ddPCR to be a more sensitive assay. Because high sensitivity is important when the amount of SARS-CoV-2 circulating in the population is low, RT-ddPCR is a valid option for the monitoring of SARS-CoV-2 in wastewater. Our study showed that isothermal amplification RT-SIBA can also be used for the detection of SARS-CoV-2 in wastewater when a qualitative result of SARS-CoV-2 is sufficient. RT-SIBA enables the detection of SARS-CoV-2 faster and with fewer resources than do RT-qPCR and RT-ddPCR.

The results of previous comparisons of RT-qPCR and RT-ddPCR have been conflicting, some finding RT-ddPCR to be superior in sensitivity, others finding RT-qPCR to be the better option, and one finding them to be similar in sensitivity. Most of these studies used different RT-qPCR and RT-ddPCR kits. As we showed in this study, there are differences in the sensitivity of a method between different kits, in our case between the TaqMan RT-qPCR assay and the QuantiTect RT-qPCR assay. When the results of different articles are compared, the reagents and methodology used should be taken into consideration.

SARS-CoV-2 GC numbers in wastewater reflect the incidence of COVID-19 in the population. Experience with SARS-CoV-2 virus monitoring has encouraged the development of monitoring methods for other human-infecting viruses in wastewater. WBS has the potential to improve the health-care system and public health preparedness for microbial epidemics and pandemics.

## Data Availability

All data produced in the present study are available upon reasonable request to the authors

## Abbreviations

GC: gene copy
WBS: wastewater-based surveillance
WWTP: wastewater treatment plant
RT-SIBA: reverse transcription strand invasion based amplification

## Ethical approval

Not required.

## Author statement

The authors have read and approved the revised version submitted.

## Acknowledgements

The authors thank Annika Laaksonen for her assistance with the laboratory work.

The authors would like to express special thanks to the personnel of the WWTPs included in the WastPan study.

The personnel of Aidian Oy, Finland, are acknowledged, with special thanks to Mirko Brummer, Antti Tullila, and Tuomas Ojalehto.

## WastPan study group

Tampere University: Sami Oikarinen, Kirsi-Maarit Lehto, Rafiqul Hyder, Annika Länsivaara and Erja Janhonen.

The Finnish Institute for Health and Welfare: Tarja Pitkänen, Ananda Tiwari, Anssi Lipponen, Anna-Maria Hokajärvi, Anniina Sarekoski, Aleksi Kolehmainen, Teemu Möttönen, Oskari Luomala, Aapo Juutinen, Soile Blomqvist, Kati Räisänen and Carita Savolainen-Kopra.

University of Helsinki: Annamari Heikinheimo, Ahmad Al-Mustapha, Viivi Heljanko, Venla Johansson and Paula Kurittu.

## Authors’ contributions

Annika Länsivaara: conceptualization, methodology, formal analysis, investigation, data curation, writing – original draft; Kirsi-Maarit Lehto: conceptualization, methodology, investigation, data curation, writing, review and editing, supervision, project administration, funding acquisition; Rafiqul Hyder: methodology, investigation, writing – review and editing; Erja Janhonen: methodology, writing – review and editing; Anssi Lipponen: writing – review and editing, project administration; Annamari Heikinheimo: writing – review and editing, project administration, funding acquisition; Tarja Pitkänen: writing – review and editing, project administration, funding acquisition; Sami Oikarinen: conceptualization, methodology, investigation, data curation, writing, review and editing, supervision, project administration, funding acquisition.

