## Supplementary material for "Comparison of Different PCR Methods for the Detection of SARS-CoV-2 RNA in Wastewater Based on the Reported Incidence of COVID-19 in Finland"

**Table 1 Used RT-qPCR and RT-ddPCR primers and probes, oligonucleotide sequences, and concentrations.**

| Name | Oligonucleotide Sequence (5'>3') | Concentration | | |
|---|---|---|---|---|
| | | TaqMan RT-qPCR | QuantiTect RT-qPCR | RT-ddPCR |
| nCoV_N1_FW | GACCCCAAAATCAGCGAAAT | 200 nM | 900 nM | 100 nM |
| nCoV_N1_RV | TCTGGTTACTGCCAGTTGAATCTG | 200 nM | 900 nM | 100 nM |
| nCoV_N1_P | FAM-ACCCCGCATTACGTTTGGTGGACC-BHQ | 200 nM | 200 nM | 25 nM |
| nCoV_N2_FW | TTACAAACATTGGCCGCAAA | 200 nM | 300 nM | - |
| nCoV_N2_RV | GCGCGACATTCCGAAGAA | 200 nM | 900 nM | - |
| nCoV_N2_P | VIC-ACAATTTGCCCCCAGCGCTTCAG-BHQ | 200 nM | 200 nM | - |
| E_Sarbeco_F | ACAGGTACGTTAATAGTTAATAGCGT | - | - | 100 nM |
| E_Sarbeco_R | ATATTGCAGCAGTACGCACACA | - | - | 100 nM |
| E_Sarbeco_P | FAM-ACACTAGCCATCCTTACTGCGCTTCG-BHQ | - | - | 25 nM |

**Table 2 TaqMan Fast Virus 1-Step RT-qPCR program**

| Step | Temperature | Time | Cycles |
|---|---|---|---|
| Reverse transcription | 50ºC | 5 min | 1 |
| RT-inactivation/initial denaturation | 95ºC | 20 s | 1 |
| Denaturation | 95ºC | 15 s | 50 |
| Annealing/extension with fluorescence data collection | 58ºC | 1 min | |

**Table 3 Qiagen QuantiTect Probe RT-qPCR program**

| Step | Temperature | Time | Cycles |
|---|---|---|---|
| Reverse transcription | 50ºC | 30 min | 1 |
| RT-inactivation/initial denaturation | 95ºC | 15 min | 1 |
| Denaturation | 94ºC | 15 s | 50 |
| Annealing/extension with fluorescence data collection | 60ºC | 1 min | |

**Table 4 The efficiency, standard curve slope, standard curve intercept, and $R^2$ of each RT-qPCR assay (N=8).**

| | TaqMan RT-qPCR N1 | | TaqMan RT-qPCR N2 | | QuantiTect RT-qPCR N1 | | QuantiTect RT-qPCR N2 | |
|---|---|---|---|---|---|---|---|---|
| | Average | Standard deviation | Average | Standard deviation | Average | Standard deviation | Average | Standard deviation |
| Efficiency | 104 | 11 | 84 | 7 | 105 | 10 | 94 | 17 |
| Std curve slope | -3,257 | 0,260 | -3,797 | 0,247 | -3,225 | 0,240 | -3,537 | 0,421 |
| Std curve intercept | 39,809 | 0,980 | 44,025 | 1,152 | 39,681 | 1,049 | 41,351 | 1,594 |
| $R^2$ | 0,983 | 0,019 | 0,990 | 0,011 | 0,983 | 0,023 | 0,984 | 0,016 |

**Table 5 Bio Rad One-Step RT-ddPCR Advanced Kit for Probes PCR program**

| Step | Temperature | Time | Cycles |
|---|---|---|---|
| | 25 | 3 min | 1 |
| Reverse transcription | 50 | 60 min | 1 |
| Enzyme activation | 95 | 10 min | 1 |
| Denaturation | 95 | 30 s | 40 |
| Annealing/extension | 55 | 1 min | |
| Enzyme deactivation | 98 | 10 min | 1 |

**Table 6 Aidian RT-SIBA program**

| Step | Temperature | Time | Cycles |
|---|---|---|---|
| Isothermal amplification with fluorescence data collection | 44ºC | 45min | 1 |
| | 95ºC | 15s | 1 |
| Melt curve with fluorescence data collection | 60ºC to 95ºC | | 1 |

**Table 7 COVID-19 incidence in each city at the sampling timepoints**

| Date | City | COVID-19 incidence/100 000 persons in sampling week |
|---|---|---|
| 22.2.2021 | Espoo | 201 |
| 22.2.2021 | Helsinki | 262 |
| 22.2.2021 | Kuopio | 11 |
| 22.2.2021 | Lappeenranta | 76 |
| 22.2.2021 | Oulu | 44 |
| 22.2.2021 | Pietarsaari | 0 |
| 22.2.2021 | Rovaniemi | 24 |
| 22.2.2021 | Seinäjoki | 11 |
| 22.2.2021 | Tampere | 37 |
| 22.2.2021 | Turku | 175 |
| 22.3.2021 | Espoo | 184 |
| 22.3.2021 | Helsinki | 234 |
| 22.3.2021 | Kuopio | 40 |
| 22.3.2021 | Lappeenranta | 103 |
| 22.3.2021 | Oulu | 11 |
| 22.3.2021 | Pietarsaari | 0 |
| 22.3.2021 | Rovaniemi | 8 |
| 22.3.2021 | Seinäjoki | 0 |
| 22.3.2021 | Tampere | 53 |
| 22.3.2021 | Turku | 235 |
| 13.12.2021 | Espoo | 296 |
| 13.12.2021 | Helsinki | 372 |
| 13.12.2021 | Kuopio | 128 |
| 13.12.2021 | Lappeenranta | 125 |
| 13.12.2021 | Oulu | 323 |
| 13.12.2021 | Pietarsaari | 36 |
| 13.12.2021 | Rovaniemi | 278 |
| 13.12.2021 | Seinäjoki | 119 |
| 13.12.2021 | Tampere | 326 |
| 13.12.2021 | Turku | 120 |
